# Prevalence and Predictors of Food Insecurity among Adolescents in Households in the Volta and Oti Regions of Ghana: A Cross-Sectional Study

**DOI:** 10.1101/2025.06.17.25328312

**Authors:** Norbert Ndaah Amuna, Deda Ogum, Juliana Yartey Enos, Timothy Yao Akweh, Israel Wuresh, Richmond Aryeetey

## Abstract

**Background:** Food insecurity poses a threat to adolescent nutrition and health, especially in low- and middle-income countries (LMICs), with potential lifelong consequences. This study assessed the prevalence and drivers of adolescent food insecurity in Ghana’s Volta and Oti regions.

**Objectives:** To assess the prevalence and predictors of food insecurity among adolescents in the Volta and Oti regions of Ghana.

**Methods:** *Design:* A community-based cross-sectional study

*Setting:* The study was conducted in the Volta and Oti regions of Ghana.

*Participants:* The sample included 667 adolescents aged 10-19 years.

*Outcome Measure:* We measured food insecurity using the Household Food Insecurity Access Scale (HFIAS). Data were collected using a two-stage sampling approach. Adolescents food insecurity status was categorised into food secure, mildly, moderately, or severely food insecure. Multivariable logistic regression analysis was employed to identify the risk factors associated with food insecurity.

**Results:** Approximately 47.5% of the surveyed adolescents reported experiencing periods of inadequate food intake during the 12 months preceding the survey. Among adolescents experiencing food scarcity, the periods from January to March (32.2%) and April to June (28.8%) were identified as the peak times of limited food availability. In total, 61% of adolescents in the Volta and Oti regions experienced food insecurity, with 23.5% classified as severely food insecure, 47.7% worried about not having enough food, and 54% could not access preferred foods due to limited resources. Approximately 50.1% consumed a monotonous diet and 44.1% consumed foods they perceived as socially unacceptable. Urban residence (aOR = 0.67; CI=0.46-0.97), higher maternal education (aOR=0.41; CI=0.22-0.78), and higher wealth (aOR=0.02; CI=0.0-0.09) were associated with reduced odds of food insecurity.

**Conclusion:** There was a high prevalence of adolescent food insecurity in the Volta and Oti regions of Ghana. This has profound implications for adolescent nutrition, health, and overall well-being. Peak periods of food scarcity align with the broader seasonal food scarcity patterns and lean seasons in Ghana. The high rates of food insecurity directly impede the goal of achieving SDG 2 and 3. Key predictors include urban-rural differences, maternal education, and wealth. Urgent action is needed to address these factors and ensure consistent access to nutritious food.

**Strengths and limitations of this study:**

- We used the widely accepted Household Food Insecurity Access Scale (HFIAS), a standardized and comprehensive tool for assessment of food insecurity.
- We employed multistage systematic sampling, enhancing representativeness and generalizability within the Volta and Oti regions.
- A cross-sectional design limits the ability to establish causal relationships.
- Potential self-reporting bias may affect data accuracy due to socially desirable or inaccurate responses.
- Findings may not be generalisable beyond the studied regions because of socioeconomic and cultural differences.

## Introduction

Food insecurity is defined as the state in which people have limited or uncertain access to sufficient, safe, and nutritious food to meet dietary needs and preferences for normal growth, development, and an active, healthy life [1–3]. It manifests at both the individual and household levels. It arises from various factors, including economic constraints, unavailability of food, insufficient purchasing power, inappropriate distribution, or inadequate use of food at the household level [1]. Food insecurity can be chronic, seasonal, or transitory, reflecting varying degrees of severity and duration [4]. Food insecurity remains a significant challenge to sustainable development, particularly in relation to Sustainable Development Goals (SDGs) one (Zero Hunger) and two (Good Health and Well-being). It disproportionately affects adolescents in low- and middle-income countries (LMICs) [5], affecting their physical, cognitive, and emotional development during a critical growth period. Insufficient nutrition during adolescence can lead to poor academic performance, increased susceptibility to infections, and a higher risk of chronic diseases in adulthood [6], undermining progress towards multiple SDGs. Furthermore, food insecurity has been linked to psychological distress, including anxiety and depression, which can affect adolescent health outcomes [7].

Globally, food insecurity is highest in resource-constrained environments, particularly in sub-Saharan Africa [5]. In Ghana, many families struggle to access sufficient and diverse foods, particularly in rural areas [8]. Food insecurity in these settings is often seasonal, with food shortages peaking during lean periods when household food stocks are depleted and market prices rise beyond the affordability of many families [9,10]. These conditions disproportionately affect adolescents, who often rely on food availability within the household and may be forced to reduce meal portions, skip meals, or consume less nutritious foods [11,12]. The Volta and Oti regions of Ghana experience high poverty rates and other environmental challenges that contribute to food insecurity. These regions are largely agrarian; many families living in these regions depend on smallholder farming for their livelihoods [13]. Agricultural productivity is influenced by factors such as erratic rainfall patterns, declining soil fertility, and limited access to modern farming inputs. Additionally, households with low income and educational levels are more likely to experience food insecurity, as economic constraints limit their ability to purchase nutritious food, leading to reliance on cheaper, calorie-dense, but nutrient-poor diets [14,15].

Existing research on food security in Ghana has primarily focused on household-level assessments targeting infants and women of reproductive age, leaving a critical gap regarding how food insecurity specifically affects adolescents. This gap is critical because adolescents have unique dietary and nutritional needs that, if unmet, increase their risk of nutritional deficiencies and hinder their physical and cognitive development [6]. Addressing adolescent food insecurity is essential to advancing SDG 2, which aims to end hunger, achieve food security, improve nutrition, and promote sustainable agriculture by 2030. Moreover, tackling adolescent food insecurity aligns with SDG 3, which focuses on ensuring healthy lives and promoting well-being for all at all ages. Therefore, understanding the prevalence and determinants of food insecurity among adolescents is critical for designing targeted interventions that enhance food access and dietary diversity in this age group, contributing to both zero hunger and improved health outcomes in Ghana.

## Methods

### Study Design

This community-based cross-sectional study was conducted in the Volta and Oti regions of Ghana. Data were collected as part of a larger study on the factors shaping adolescents’ diet diversity, food security, reactions to advertising, nutritional status, and readiness to adopt sustainable healthy diets.

### Study Settings and Period

This study was conducted in forty-seven communities located in twenty-six districts and municipalities in the Volta and Oti regions of Ghana. Data were gathered from August 2021 to March 2022. The Volta and Oti regions are two of the sixteen administrative regions in Ghana; they cover a total land mass of 20,572 Km^2^, representing 8.7 percent of the total land area of Ghana. The area extends northward from the Gulf of Guinea, traversing all of Ghana’s major vegetation zones, including tropical rainforest, moist semi-deciduous forest, coastal scrub and grassland, strand and mangrove, and Guinea savannah. It is bordered to the west by Lake Volta, to the east by the Republic of Togo, and to the north by the Northern Region. The region’s primary economic activities are agriculture, fishing, and trading, reflecting the diverse natural resources and ecological conditions across its landscape.

### Participants

The study participants were adolescents aged 10-19 years. A representative sample of 667 adolescents was obtained from both urban and rural areas within the Volta and Oti regions, using a multi-stage random sampling method.

### Sample Size Determination

The minimum sample size was calculated using Yamane’s formula for population proportion, n=N/(1+Ne^2^) [16], where n represents the minimum sample size, e is the margin of error set at 5%, and N is the projected adolescent population in the two regions, which was estimated to be 589,513 in 2020. The calculated sample was multiplied by a design effect of 1.5, as a multi-stage sampling technique was used, and a 10% non-response rate was added. The total estimated sample size was 667.

### Sampling Technique and Procedures

This study employed a two-stage stratified sampling design. Enumeration areas (EAs), based on those used for the 2017 Ghana Living Standards Survey round 7 (GLSS 7), were selected as the Primary Sampling Units (PSUs). Forty-five (45) EAs from a total of ninety-six were chosen to form the PSUs. These PSUs were allocated to the two administrative regions using probability proportional to population size (PPS) and further divided into urban and rural localities of residence. In the second stage, Secondary Sampling Units (SSUs) were identified using a complete listing of households in the selected PSUs. The total sample size was 675 adolescents in the two regions. Fifteen households from each PSU were systematically selected. Within each selected PSU, a comprehensive listing of households was compiled to serve as the Secondary Sampling Units (SSUs). From these household lists, systematic sampling was used to select 15 households per PSU. The sampling interval (k) was determined by dividing the total number of households in the PSU by 15. For instance, if a PSU comprised 150 households, every 10th household was selected. A random starting point was identified using landmarks to initiate the selection process, thereby ensuring that each household had an equal and known chance of inclusion. This approach ensured the spread of sampled households throughout the PSU, thereby minimising clustering and enhancing representativeness.

### Inclusion and Exclusion Criteria

Adolescents between the ages of 10 and 19 years, residents in the Volta or Oti regions, and willing to voluntarily participate in the study were considered eligible for this study. Adolescents who met the inclusion criteria but were critically ill or had cognitive or physical disabilities impacting their ability to participate, institutionalisation (e.g. prison, hospitalisation), and adolescents who had insufficient linguistic fluency to complete the study procedures/questionnaires were excluded.

### Data Collection Tool and Procedures

Structured, pre-tested questionnaires were administered face-to-face to the participants to collect data. These questionnaires were designed to capture sociodemographic characteristics and food security measures in order to contextualise food insecurity findings. The data collection process employed the Kobo Toolbox, an open-source digital mobile platform that enables real-time data entry, monitoring, and immediate quality checks. A research team comprising 15 experienced nutrition and public health graduates conducted data collection under the supervision of three nutrition professionals. The interviews were conducted in participants’ homes and other community spaces where participants’ and interviewers’ privacy, confidentiality, and safety could be maximised. The questions were asked in local Ghanaian languages or English.

### Variables and Measures

#### Food security status

Food insecurity status (FIS) was assessed using the Household Food Insecurity Access Scale (HFIAS) developed by the USAID-funded Food and Nutrition Technical Assistance (FANTA) Project in 2006 [17]. The HFIAS has been previously validated in multiple African contexts, including Burkina Faso [18] and Tanzania [19].The HFIAS tool consists of nine core questions that capture households’ experiences related to food access within the previous four weeks. This methodological approach is founded on the understanding that food insecurity experiences trigger predictable reactions and responses that can be systematically measured and quantified using standardised survey instruments. The nine questions in the HFIAS assess multiple dimensions of food insecurity.

1. Anxiety about household food supply
2. Insufficient food quality (including variety and preferences)
3. Insufficient food intake and its physical consequences
4. Socially unacceptable means of food acquisition

Each of the nine questions was asked with reference to the four weeks (30 days) preceding the survey. Responses were coded on a frequency-of-occurrence basis using a 3-point Likert scale: 0 = Never; 1 = Rarely (1-2 times); 2 = Sometimes (3-10 times); 3 = Often (more than 10 times)

The Food Insecurity Access Scale (FIAS) score was calculated as a continuous measure by summing the coded responses across all nine questions. The resulting scores ranged from 0 (complete food security) to 27 (severe food insecurity). Based on the individual FIAS scores, adolescents were classified into four food security categories according to the HFIAS guidelines: 0-1 = Food secure; 2-8 = Mildly food insecure; 9-16 = Moderately food insecure; and 17-27 = Severely food insecure [17]. The internal consistency reliability of the HFIAS was confirmed with a Cronbach’s alpha coefficient of 0.72, indicating acceptable reliability for the study population. This categorisation process excluded responses related to voluntary fasting or dietary restrictions for weight loss to ensure that the measurement captured only involuntary food restriction due to resource constraints.

### Data Quality Control

To minimise bias, standardised tools were used to measure food insecurity and other variables. Trained interviewers collected the data. Selection bias was minimised through systematic sampling. Reporting bias was reduced through questionnaire validation. A pre-test was conducted on eligible adolescents in Kpeve and Kete Krachi, comparable settings that were not involved in the research. The administered questionnaires were checked for completeness daily before being collected from the data collectors.

### Data Processing and Analysis

Data were entered using EpiData and exported to STATA version 17 software for analysis. Sociodemographic characteristics were considered independent variables. Frequencies, means, and percentages were used to summarise the results. Logistic regression was used to identify predictors of food insecurity. Bivariate and Multivariate logistic regression models were used to analyse the data. The significance level was set at P ≤ 0·05. All analyses were conducted using Stata software [20].

## Results

### Background Characteristics of the Population

Table 1 shows the background characteristics of the study population. A total of 667 adolescents aged 10-19 years were surveyed (98.8% of the sample). The majority (70.2%) of the adolescents were from the Volta region, and 63.3% lived in rural areas. Females accounted for 59.5% of the sample. The mean age of the adolescents was 15.1±2.5 years, with 43.2% aged 15-17 and 36.7% aged 10–14 years. The study participants were predominantly Christian (85.9%) and of Ewe ethnicity (65.1%). Approximately, 43% had five or more siblings. In terms of education, the majority (632, 94.8%) were enrolled in school, 41.4% in Junior High School (JHS), and 35.5% in Senior High School (SHS). Regarding school type, 82.7% attended public schools. Family structure analysis showed that 94.6% of participants had never married and 5.4% were married or co-habiting. Regarding living arrangements, 81.3% lived with both parents. The occupational status of mothers showed that 48.9% were primarily traders; 34.6% were professionals, such as doctors, nurses, teachers, midwives, and head teachers; and 33.7% were farmers (33.7%). In terms of maternal education, 26.5% had basic education and 24.3% had completed Junior High School. The average household size was 6.3±2.8 members. In terms of wealth distribution, 38.7% of households were in the fourth quintile and 32.4% in the fifth quintile.

**Table 1:**
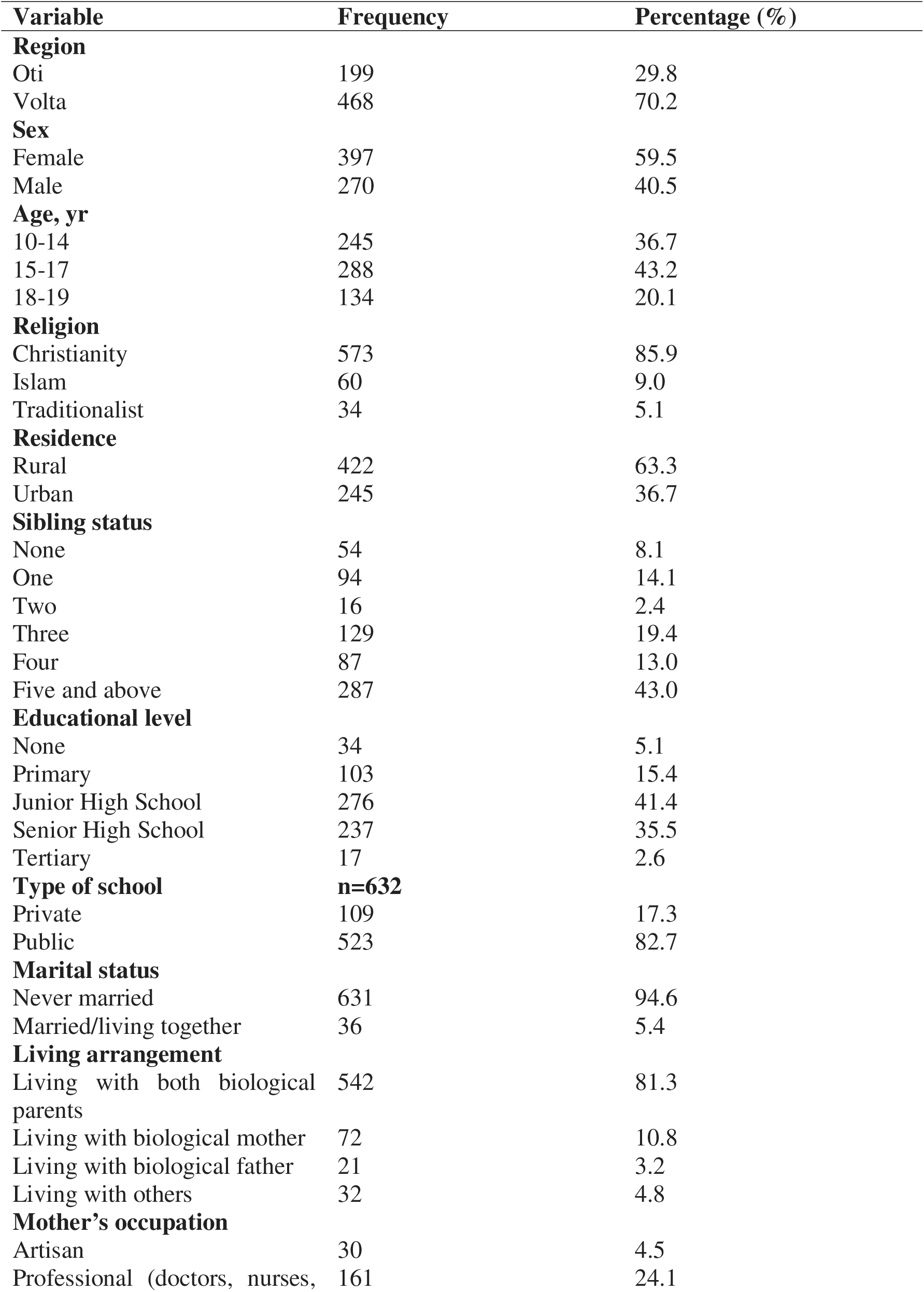
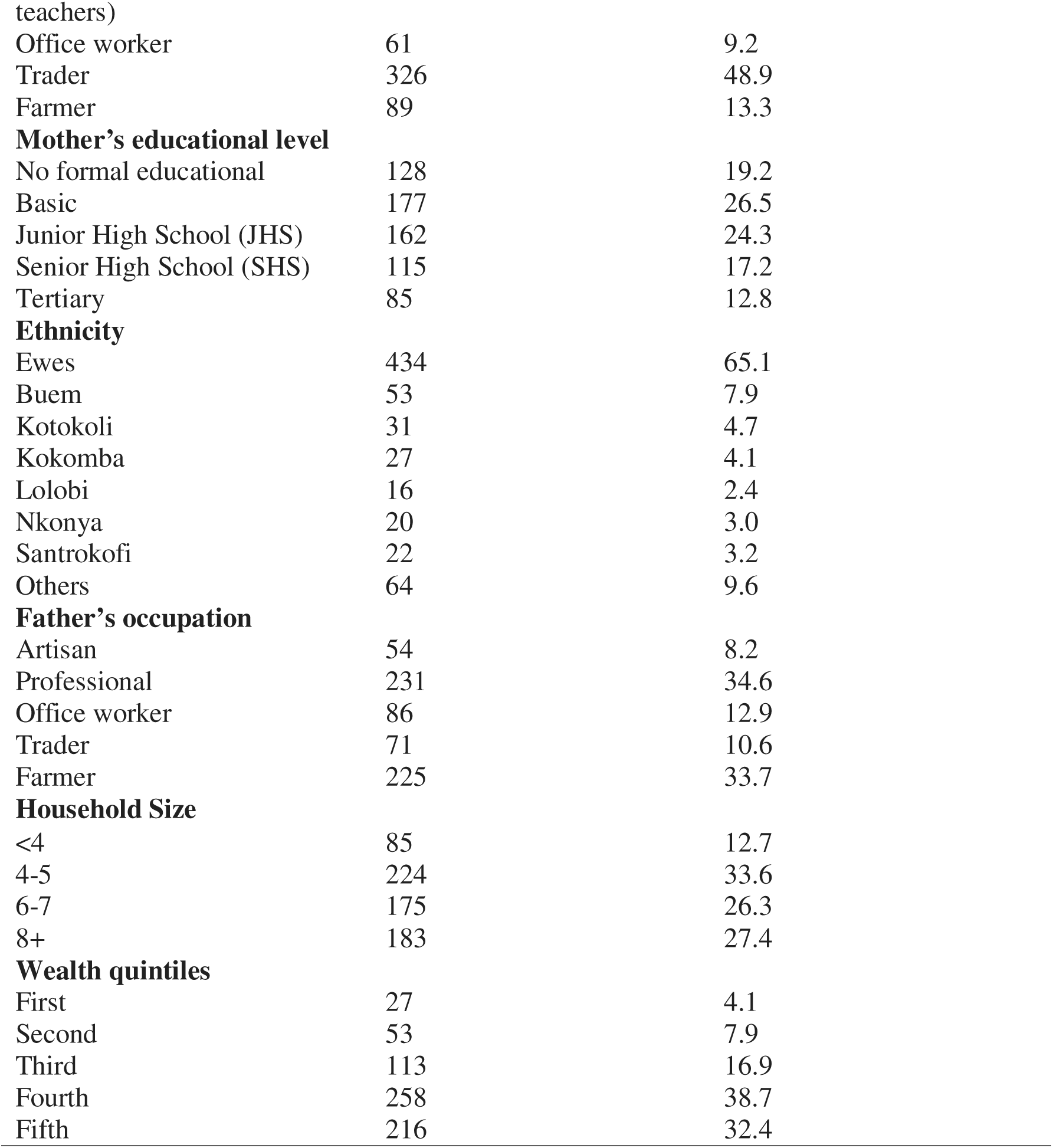
Background Characteristics of Adolescents (n=667)

### Prevalence and levels of food insecurity among adolescents

Table 2 shows the results of the food insecurity analysis among adolescents in the Volta and Oti regions of Ghana. Nearly half of the surveyed respondents (47.5%) reported that there were months in the past year during which they did not have enough food to meet their families’ needs. The period of peak food scarcity was observed from January to March, during which 32.2% of the adolescents reported experiencing food shortages, followed by April to June, with 28.8% of the adolescents indicating similar challenges. A declining trend in food scarcity was noted, as both the number and percentage of families facing food shortages consistently decreased in each subsequent quarter of the year. The survey results showed that 32.8% of adolescents had enough of their preferred foods, while 28.7% had sufficient food supply, but with limited variety or options that aligned with their preferences. However, 13.8% of respondents indicated that they occasionally experienced food insufficiency, and 24.7% reported that they frequently experienced food insufficiency.

**Table 2:** Food Insecurity among Adolescents.

| Variable | Food security status |  |
| --- | --- | --- |
|  | Frequency | Percentage |
| Over the last 12 months there were months when there was not enough food to meet needs of family | 317 | 47.5 |
| In the past 12 months, months family did not have enough food to meet needs. |  |  |
| *January-March* | 102 | 32.2 |
| *April-June* | 91 | 28.8 |
| *July-September* | 74 | 23.3 |
| *October-December* | 50 | 15.6 |
| Description of the food eaten in your household in the last 12 month. |  |  |
| *Enough of the kinds of food we want to eat* | 219 | 32.8 |
| *Enough but not always the kinds of food we want* | 191 | 28.7 |
| *Sometimes not enough to eat* | 92 | 13.8 |
| *Often not enough to eat* | 165 | 24.7 |
| Worried there would not be enough food to eat in the last four weeks | 318 | 47.7 |
| Unable to eat preferred food in the last four weeks due to lack of resources. | 360 | 54.0 |
| Ate a limited variety of foods (monotonous diet) | 334 | 50.1 |
| Ate undesirable food or food I didn't want (socially unacceptable food). | 294 | 44.1 |
| Cut the size of meals or ate smaller meals | 444 | 66.6 |
| Ate fewer meals in a day. | 232 | 34.8 |
| Had no food to eat of any kind (run out of food). | 221 | 33.2 |
| Went without eating a whole day and night. | 150 | 22.5 |
| Went to sleep at night without eating | 213 | 31.9 |
| Food security status |  |  |
| *Food secure* | 260 | 39.0 |
| *Food insecure* | 407 | 61.0 |
| *Mildly food insecure* | 147 | 22.0 |
| *Moderately food insecure* | 103 | 15.4 |
| *Severely food insecure* | 157 | 23.5 |

Almost half (47.7%) of the adolescents surveyed reported being worried or concerned that they would not have enough food to eat or sufficient food to meet their needs, and 54% could not consume or afford their preferred food due to resource constraints. Adolescents resorted to using various strategies to cope with food insecurity. This included eating a limited variety of foods (monotonous diet) (50.1%) and eating foods that they did not want or found socially unacceptable because they lacked the resources to acquire other types of food (44.1%). Two-thirds of adolescents (66.6%) reported having reduced the size of meals, and 34.8% reported eating fewer meals (skipping meals) each day because there was not enough food. The results reveal an alarming severity of food insecurity among adolescents: one-third (33%) of adolescents faced complete depletion of food resources in their households, leaving them entirely without sustenance due to financial constraints. Even more disturbingly, nearly a quarter (22.5%) of adolescents endured extreme hunger, going through an entire 24-hour period without a single meal due to a lack of food or financial means. Overall, 61% of adolescents were food insecure, and these were either mild (22.0%), moderate (15.4%), or severe (23.5%).

### Factors associated with adolescent food insecurity

The findings presented in Table 3 suggest that regional disparities are associated with food insecurity. Adolescents in the Volta region had higher odds of being food insecure compared to the Oti region (aOR = 1.61; CI=1.09-2.39). Urban residence was associated with reduced odds of food insecurity, with urban dwellers having 67% less odds of experiencing food insecurity compared to rural areas (aOR = 0.67; CI=0.46-0.97). Higher maternal education level was associated with lower odds of food insecurity; mothers who had completed Junior High School (aOR=0.41; CI=0.22-0.78) or Senior High School (aOR=0.38; CI=0.19-0.75) had significantly lower odds of food insecurity than those with no formal education. Additionally, higher wealth quintiles were significantly associated with lower odds of food insecurity, with the second (aOR=0.15; CI=0.03-0.67), third (aOR=0.05; CI=0.01-0.21), and fourth (aOR=0.02; CI=0.0-0.09) wealth quintiles showing lower odds of food insecurity. These findings highlight the importance of parental education and economic status in determining food security.

**Table 3:**
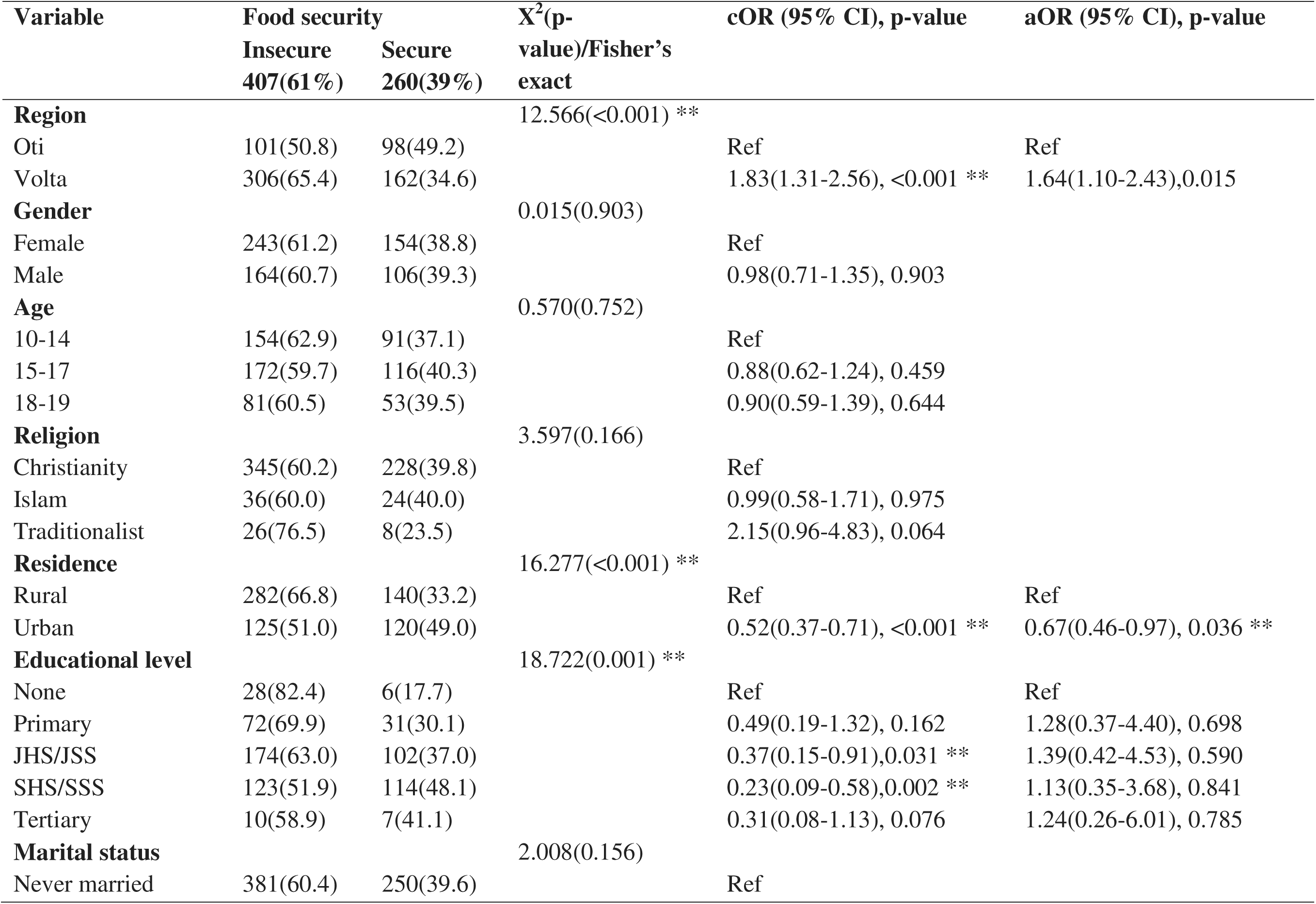
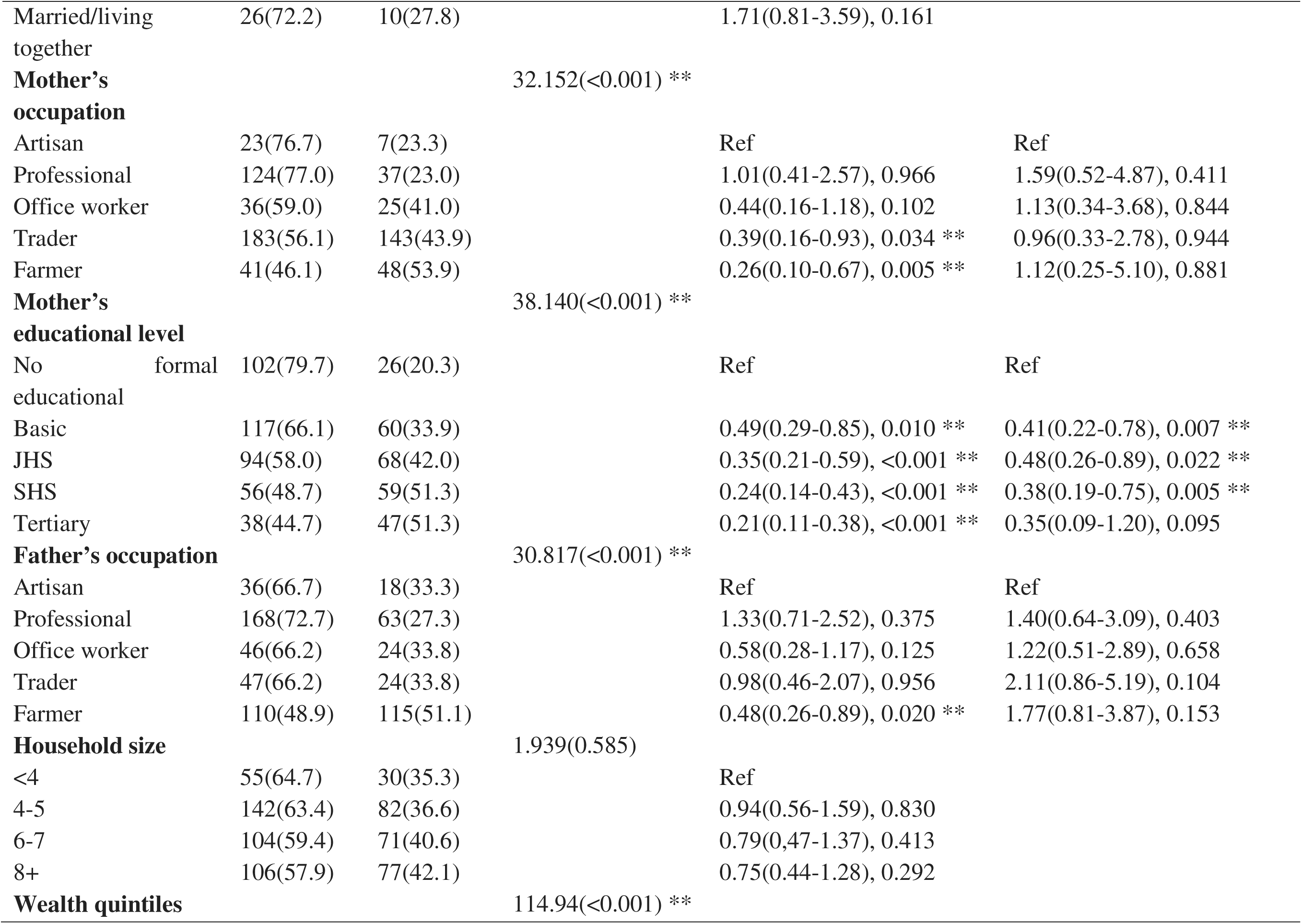
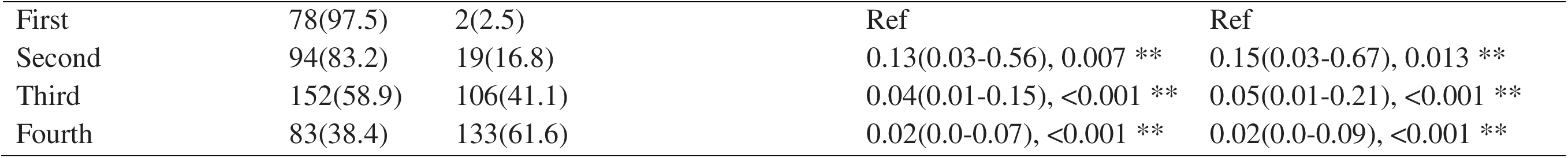
Food security and its associated factors.

## Discussion

There is a high prevalence of food insecurity among adolescents in the Volta and Oti regions of Ghana, with 61% of adolescents experiencing food insecurity ranging from mild to severe. This level of food insecurity poses serious risks to adolescent health; cognitive, physical, and socio-emotional development; and overall well-being. This situation is critical, requiring an urgent need for targeted interventions to address this issue. As a stage of rapid physical and cognitive development, adolescents need adequate nutritional intake, making food shortages particularly detrimental [6,7]. Food insecurity is linked to poor health outcomes, including nutritional deficiencies and mental health issues [21]. Studies worldwide have highlighted the lasting effects of food insecurity at the adolescent stage. Malnutrition, resulting from inadequate food intake, impairs physical and cognitive development, contributes to stunting, reduces productivity, increases psychological distress, and increases susceptibility to diseases [22,23]. The persistence of food insecurity in these regions reflects broader global trends, where adolescents in low-income settings face substantial risks owing to their unique nutritional vulnerabilities [24].

This study found that food scarcity persisted for several months, peaking during the first two quarters of the year (January to June), with the most pronounced peak occurring in the first quarter of the year. These peak periods of food scarcity align with the lean season in Ghana, the time before the main harvest, which is characterised by diminished food availability and increased food prices, exacerbating food insecurity for many households [25,26]. This trend is consistent with patterns observed across sub-Saharan Africa, where seasonal fluctuations in food availability significantly impact household food security [27–29]. This observation suggests a seasonal effect, necessitating the implementation of support mechanisms to alleviate recurrent shortages. In rural Ghana, where subsistence farming is the primary livelihood, this dependence on unpredictable harvests exacerbates food insecurity [13]. Households that rely on their own agricultural output for both consumption and income are particularly vulnerable, highlighting the need for targeted interventions such as food assistance or financial support during critical periods.

Adolescents facing food scarcity resorted to adopting various unhealthy coping strategies, such as consuming monotonous diets or nutritionally inadequate foods, skipping meals, and reducing meal sizes. The extreme cases of complete food depletion and 24-hour periods without meals for a significant portion of adolescents highlight the critical nature of the situation, potentially impacting their physical and cognitive development, academic performance, and socioeconomic prospects. Similar behaviours have been observed in other low-income regions, where survival often takes precedence over dietary quality [22,30]. Unhealthy coping strategies have serious consequences. Diets lacking diversity increase the risk of micronutrient deficiencies, while prolonged hunger impairs cognitive function and academic performance [23,31]. Such coping strategies may also lead to long-term health consequences, with the possibility of perpetuating intergenerational cycles of malnutrition, especially for female adolescents [32,33]. The psychological burden is also significant, as food insecurity has been linked to heightened stress and anxiety among adolescents [34]. The emotional strain of uncertainty about food availability further compounds the adverse effects, reinforcing cycles of poor mental and physical health [23].

Food insecurity is not solely an issue related to agricultural production or food availability. It is deeply rooted in the socioeconomic conditions that shape people’s access to resources, opportunities, and entitlements. Various factors contribute to the availability, accessibility, utilisation, and stability of food systems [35]. In this study, the predictors of adolescent food insecurity were mainly socioeconomic factors. Wealthier households generally experience lower levels of food insecurity. This reflects the broader correlation between poverty and hunger, where limited financial resources restrict access to sufficient and nutritious food [36,37]. Addressing this requires economic empowerment programs and social safety nets to support vulnerable households. Maternal education has emerged as a critical determinant, with adolescents from more educated mothers being less likely to experience food insecurity. This finding is consistent with global research demonstrating that educated mothers are better equipped to manage household resources and make informed nutritional choices [38]. Higher maternal education is associated with better home food environments and dietary practices [39]. Therefore, promoting female education serves as a long-term strategy to enhance household food security and overall well-being. Economic determinants, including income levels, employment opportunities, and livelihood diversification, play a pivotal role in determining households’ ability to acquire sufficient, nutritious food.

Urban residency was associated with decreased odds of experiencing food insecurity. Households in urban areas typically engage in cash-based economies and are more likely to benefit from stable formal employment and social safety nets, which can alleviate food insecurity [40,41]. In contrast, rural populations often face seasonal income fluctuations and climate-related agricultural risks [41]. Urban regions generally offer better access to markets with diverse food options [41,42], as well as water and electricity services essential for food preservation and storage [42,43] and transportation networks that facilitate food distribution. In contrast, rural areas often lack these amenities, exacerbating challenges related to food access. Many rural areas in low- and middle-income countries (LMICs) lack food retailers that provide fresh and affordable foods. This scarcity is often attributed to small population sizes, low-income levels, and economic challenges that render it unprofitable for supermarkets or large grocery stores to operate sustainably. Consequently, rural residents depend on small convenience stores or distant markets that often offer limited, less nutritious, and more expensive food options. Similar disparities are observed globally, where geographical location significantly affects access to food and economic opportunities [44]. Addressing these inequalities requires targeted policies aimed at enhancing infrastructure, economic investment, and resource allocation in underserved areas. Urbanisation adds further complexity, as urban adolescents generally experience less severe food insecurity but face economic constraints due to higher living costs [45]. Although urban households may have better access to markets and social services, financial strain remains a significant barrier to food security. The long-term consequences of adolescent food insecurity cannot be overlooked. Poor nutrition at this stage has lifelong implications, as it limits physical development, reduces cognitive capacity, and hinders economic productivity. Research highlights that malnutrition during adolescence not only affects individual health outcomes but also weakens national development by diminishing human capital potential. Adolescents who grow up in food insecure environments are less likely to complete their education or achieve economic stability, thereby perpetuating cycles of poverty and inequality.

## Conclusion

Food insecurity affects over 60% of adolescents in Ghana’s Volta and Oti regions, with 25% experiencing severe food insecurity. This situation adversely impacts nutrition, health, and cognitive development, with seasonal vulnerability exacerbating the issue. High rates of food insecurity, especially severe food insecurity, directly impede the goal of ending hunger and ensuring access to safe, nutritious, and sufficient food for all (SDG 2), and the goal of ensuring good health and well-being (SDG 3). The consequences are profound, both in the short and long term, affecting health, development, and intergenerational outcomes, including malnourished mothers and low-birth-weight infants. Addressing this challenge requires integrated, context-specific strategies that extend beyond mere food provision to address the underlying socioeconomic determinants. Such efforts are essential for achieving sustainable development and safeguarding the health and well-being of Ghana’s youth and future generations. Protective factors include urban residence, higher maternal education, and improved socioeconomic status. Comprehensive strategies are required to address immediate food needs and underlying socioeconomic factors. Interventions should focus on enhancing food access, nutrition education, and reducing socioeconomic disparities. Collaboration among stakeholders is crucial for the sustainable development and well-being of current and future generations.

## Article Summary

### Strength and Limitations of the Study

This study exhibits methodological robustness through the adoption of the widely accepted Household Food Insecurity Access Scale (HFIAS), ensuring a standardised and comprehensive evaluation of food insecurity. The multistage random sampling methodology enhances the study’s representativeness, allowing for generalisability to the broader population in the Volta and Oti regions. While this study employed a robust design, its cross-sectional nature restricted the establishment of causal relationships. In addition, self-reporting bias may affect the accuracy of the data, as respondents may provide socially desirable or inaccurate information about their food security experiences. The results may be applicable to other rural and urban Ghanaian settings; Generalisability in other LMICs would require further study because of socioeconomic and cultural differences.

## Data Availability

All data produced in the present study are available upon reasonable request to the authors

## Declarations

This study was conducted without external funding support. All research activities were self-funded by the authors.

### Author Contributions

Conceptualisation: NDA, RA, DO Methodology: NDA, RA; Formal analysis: NDA, TYA, IW; Writing - original draft: NDA, IW, TYA; Writing - review and editing: DO, JYE, IW, RA; Project administration: NDA, TYA; Supervision: RA, JYE, DO. All authors have read and agreed to the published version of the manuscript

### Ethical Approval

Ethical approval for this study was obtained from the Institutional Review Board of Noguchi Memorial Institute for Medical Research (NMIMR-IRB), University of Ghana (NMIMR-IRB CPN 051/20-21).

### Informed Consent Statement

Informed consent was obtained from all participants prior to inclusion in the study. However, for participants less than 18 years old, written parental consent and child assent were obtained. All ethical guidelines regarding the use of human participants were strictly adhered to in this study.

### Consent for publication

Written informed consent was obtained from the participant(s) to publish the results of the study.

### Data Availability Statement

Data are available upon request from the corresponding author.

## Acknowledgements

Mr. Martin Adjuik provided guidance on study design and statistical analysis. The authors acknowledge the participants of this study.

## Patients and the public involvement

Patients and the public were not involved in this study.

